# Rapid non-invasive detection of Influenza-A-infection by multicapillary column coupled ion mobility spectrometry

**DOI:** 10.1101/2020.06.04.20099259

**Authors:** Claus Steppert, Isabel Steppert, Thomas Bollinger, William Sterlacci

## Abstract

Infectious pathogens are a global issue. Global air travelling offers an easy and fast opportunity not only for people but also for infectious diseases to spread around the world within a few days. Also, large public events facilitate increasing infection numbers. Therefore, a rapid on-site screening for infected people is urgently needed.

Due to the small size and easy handling, the ion mobility spectrometry coupled with a multicapillary column (MCC-IMS) is a very promising, sensitive method for the on-site identification of infectious pathogens based on scents, representing volatile organic compounds (VOCs).

The purpose of this study was to prospectively assess whether identification of Influenza-A-infection based on VOCs by MCC-IMS is possible in breath. Nasal breath was investigated in 24 consecutive persons with and without Influenza-A-infection by MCC-IMS. In 14 Influenza-A-infected patients, infection was proven by PCR of nasopharyngeal swabs. Four healthy staff members and six patients with negative PCR result served as controls. For picking up relevant VOCs in MCC-IMS spectra, software based on cluster analysis followed by multivariate statistical analysis was applied. With only four VOCs canonical discriminant analysis was able to distinguish Influenza-A-infected patients from not infected with 100% sensitivity and 100% specificity.

This present proof-of-concept-study yields encouraging results showing a rapid diagnosis of viral infections in nasal breath within 5 minutes by MCC-IMS. The next step is to validate the results with a greater number of patients with Influenza-A-infection as well as other viral diseases, especially COVID-19.

Registration number at ClinicalTrials.gov NCT04282135.

## 1. Introduction

Air travel not only leads to global connection of people and goods but also to an easy spread of infectious diseases [1]. During the SARS epidemic non-contact temperature measurements were used to recognize infected passengers before boarding but even for the detection of fever there is a lack of specificity and sensitivity [2].

The need for mass screening of viral diseases is highlighted in the current COVID-19 pandemic, when even most developed countries’ health services operate under enormous strain.

So, a rapid detection of communicable viral diseases is urgently needed not only to avoid spread by air travel but also to detect and cut infection chains.

Ancient Greek physicians already used their sense of smell to detect diseases, especially infectious diseases [3]. These scents are volatile organic compounds (VOC) released during the metabolism of the bacteria or the host. VOCs are organic chemical compounds that are typically present in the gas-phase under ambient conditions of temperature and pressure

Different analytic procedures including multicapillary column coupled ion mobility spectrometry (MCC-IMS) can be used to differentiate between bacterial species in vitro [4,5]. A detection of bacterial infection could also be achieved in the breath of infected animals as well as in tuberculosis-infected humans [6,7]. Unlike bacteria, viruses do not have a metabolism of their own but VOCs can also be detected when produced by the infected host cells or as a result of immunologic defense mechanisms [8].

The present study was designed as a proof-of-concept to show whether it is possible to detect Influenza-A-infections by MCC-IMS of exhaled breath.

## 2. Methods

### 2.1 Patients

Eligible patients included male and female adults (aged ≥ 18 years) that were admitted to our hospital as well as four asymptomatic staff members. According to hospital policies during the Influenza season in every patient a nasopharyngeal swab was taken once at admission for the PCR-analysis for Influenza RNA. Patients were consecutively asked to participate in the study.

The study was approved by the ethics committee of the university of Erlangen (Nr 426_18 B) and conducted according to the principles of the Declaration of Helsinki, good clinical practice and the European General data protection regulation. All patients gave written informed consent prior to participation. The study is registered at ClinicalTrials.gov (NCT04282135).

### 2.2 Polymerase Chain Reaction (PCR)

Influenza-A was tested by taking a deep nasopharyngeal swab applying the “Xpert®Nasopharyngeal Sample Collection Kit for Viruses” (Cepheid, Maurens-Scopont, France) and performing Influenza-A PCR with the “Xpert® Xpress Flu/RSV” (Cepheid) on the “Infinity” (Cepheid). The negative predictive value of this test is reported to be between 99.7 - 100%. [9].

### 2.3 Breath sampling and IMS analysis

As the primary replication site of the Influenza-A virus is the nasopharyngeal mucosa, nasal breath was aspirated for 10 seconds during normal respiration by a foam cuffed oxygen catheter (#01442958, Asid-Bonz, Herrenberg, Germany) connected via a 0.22μm Filter (Navigator Lab Instruments, Tinajin, China) and a Perfusor Line (B Braun, Melsungen, Germany) to the MCC-IMS and directly analyzed without any pre-analytic procedures. Every patient was examined by this procedure in the first 24h after admission to ensure enough virus loads in the upper airways.

For the VOC analysis a MCC-IMS-device from STEP Sensortechnik und Elektronik, Pockau, Germany (STEP IMS NOO) was used. The breath sample was drawn into the sampling loop using an internal pump with a flow rate of 200 ml/min. In the sampling loop the collected sample was pre-separated by isothermally heated multicapillary gas chromatography column (60°C) into single analytes, which finally entered the IMS unit based on their retention times. In the IMS unit the analytes were ionized by beta radiation with a tritium source (99 MBq). Afterwards the generated ions were accelerated in a 50mm-long drift tube under the influence of an electric field (400 V/cm) towards the detector which is also tempered to 60°C. On their drift way they collide with air molecules from the drift gas (400 ml/min) flowing in the opposite direction getting separated depending on their ion mobilities and detected by the collector electrode sampled every 10μs. IMS measurements were performed in positive ionization mode. The received IMS spectra were stored internally in the device and later analyzed offline.

The used IMS device is equipped with a circulation filter and internal gas circulation. Using a circulation pump, ambient air filtered by activated carbon was provided as drift gas and analysis gas (20 ml/min) to the device. For the IMS- settings see web-only Supplementary Table S1.

### 2.4 IMS data analysis

For a total of 240s one spectrum was derived every second of retention time and a drift time of 0 to 20.48ms. As the VOC are physically determined by their retention time and the drift time heatmaps were generated with retention time on the Y-axis and drift time on the X-axis.

These heatmaps were further analyzed by proprietary cluster analysis software using support vector machine [6,10]. After baseline correction for noise the software determines the clusters based on the signal threshold and categorizes them by retention time and drift time. Depending on these parameters the clusters are numbered (see web-only Supplementary Table S2).

### 2.5 Statistical analysis

For the demographics and laboratory findings descriptive statistics were used. Due to the high cross-correlation of the clusters multivariate analysis had to be employed. We decided to perform a stepwise canonical discriminant analysis for the classifier. Signal intensities in Volt [V| of the clusters as variables were considered in statistical analysis. The discriminant analysis was aimed for a maximal reduction of Wilks-Lambda with maximum significance of F to enter a variable of 0.05 and a minimum significance to remove of 0.1. Cross-validation was performed by the leave-one-out method.

For the statistical analysis we used IBM SPSS 26.0 (IBM, Armonk, NY).

## 3. Results

### 3.1 Patient characteristics

Between February 15^th^ 2020 and March 5^th^ 2020 4 healthy staff members and 20 patients with suspected Influenza-A-infection and PCR confirmed Influenza-A status were included in our study. For details please refer to table 1.

**Table 1.** Demographics of included patients including leukocyte count C-reactive Protein (CRP) and procalcitonine (PCT) as inflammation markers

| Demographics | Influenza-A<br>negative | Influenza-A<br>positive | Total |
| --- | --- | --- | --- |
| Male/Female | 7/3 | 4/10 | 11/13 |
| Age (yrs) | 61.6 ± 14.8 | 69.9 ± 13.2 | 66.4 ± 14.2 |
| Leukocytes [G/L] | 7.3 ± 3.7<br>1.5 - 11.5 | 7.8 ± 2.7<br>2.2 - 12.1 | 7.6 ± 2.9<br>1.5 - 12.1 |
| CRP [mg/L] | 38.5 ± 28<br>4-66 | 76 ± 64<br>1 -240 | 65 ± 57<br>1 -240 |
| PCT [ng/L] | 2.2 ± 1.8<br>1-5.6 | 3.7 ± 9.1<br>0.7-35.4 | 3.3 ± 7.6<br>0.7 - 35.4 |

### 3.2 MCC-IMS

Using cluster analysis software, a total of 1033 clusters were found. Clusters where the median signal intensity of Influenza-A negatives exceeded the median signal intensity of Influenza-A positives were omitted, resulting in 74 usable clusters.

Using stepwise canonical discrimination analysis 7 clusters could optimally minimize Wilks Lambda to 0.062. Despite an imbalance of gender between infected and non-infected subjects gender had no influence on discrimination in the multivariate analysis. Using these 7 clusters a sensitivity of 100% and a specificity of 100% could be found at cross-validation by leave-one-out (see web-only Supplementary Table S3 and S4).

Additionally, another 3 clusters could be removed maintaining 100% specificity and 100% sensitivity at cross validation of the remaining 4 clusters (Table 2). In Figure 1 the peak intensities of these 4 clusters are depicted.

**Table 2.**
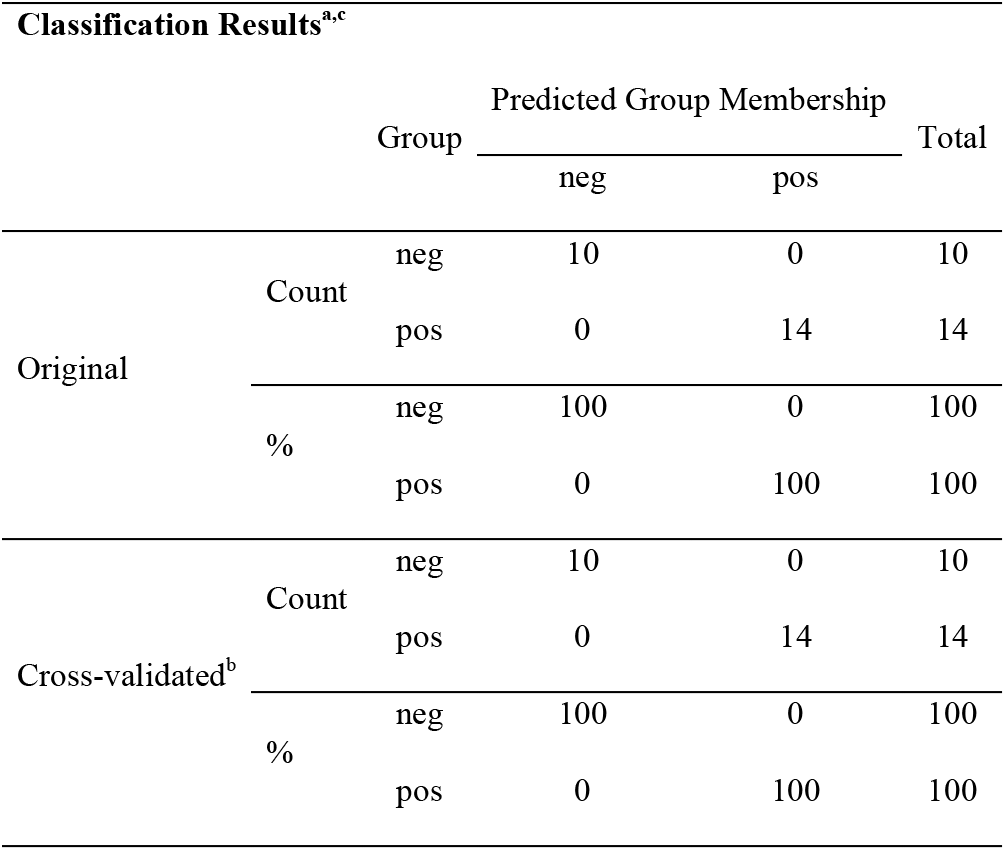
Results of linear canonical discrimination analysis using 4 clusters c_521 (164s, 4.41ms), c_378 (119s, 4,56ms), c_345 ((117s, 4,25ms) and c_116 (44s, 2,69ms). Neg = Influenza-A negative, pos = Influenza-A positive. a. 100,0% of original grouped cases correctly classified. b. Cross validation is done only for those cases in the analysis. In cross validation, each case is classified by the functions derived from all cases other than that case. c. 100% of cross-validated grouped cases correctly classified.

**Figure 1.**
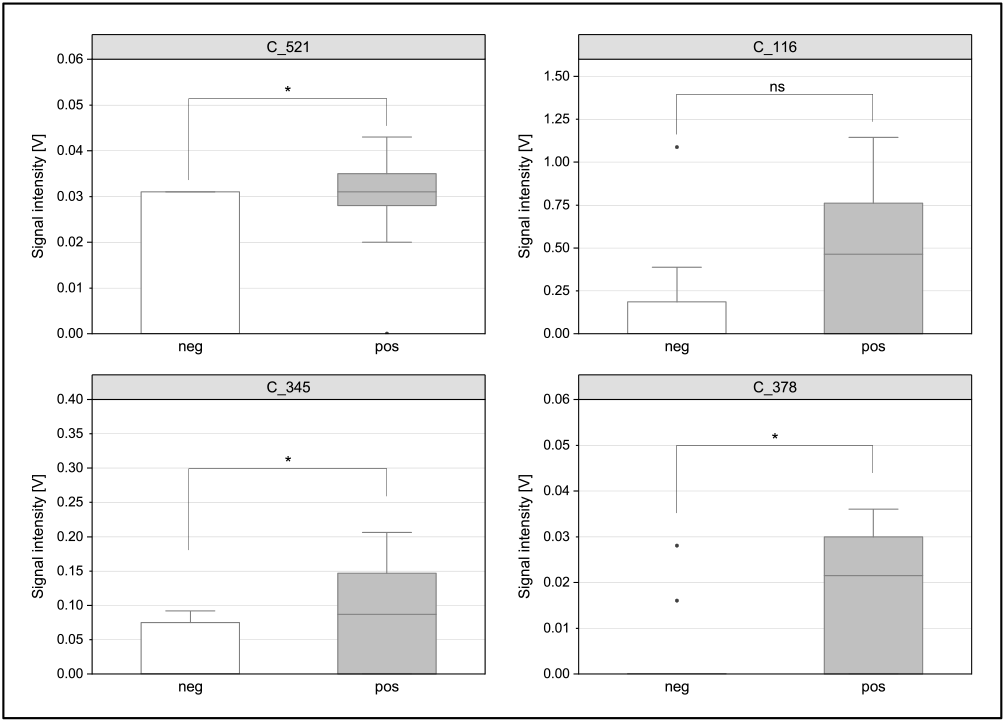
Boxplots of the peak intensities for the 4 most significant clusters C_116, C_345, C_378, C_521 in the multivariate analysis.

Fig. 2 shows the separation of the positive and negative patients in a 3D-plot of the signal intensities of the 3 most significant clusters: c_116, c_378 and c_521.

**Figure 2.**
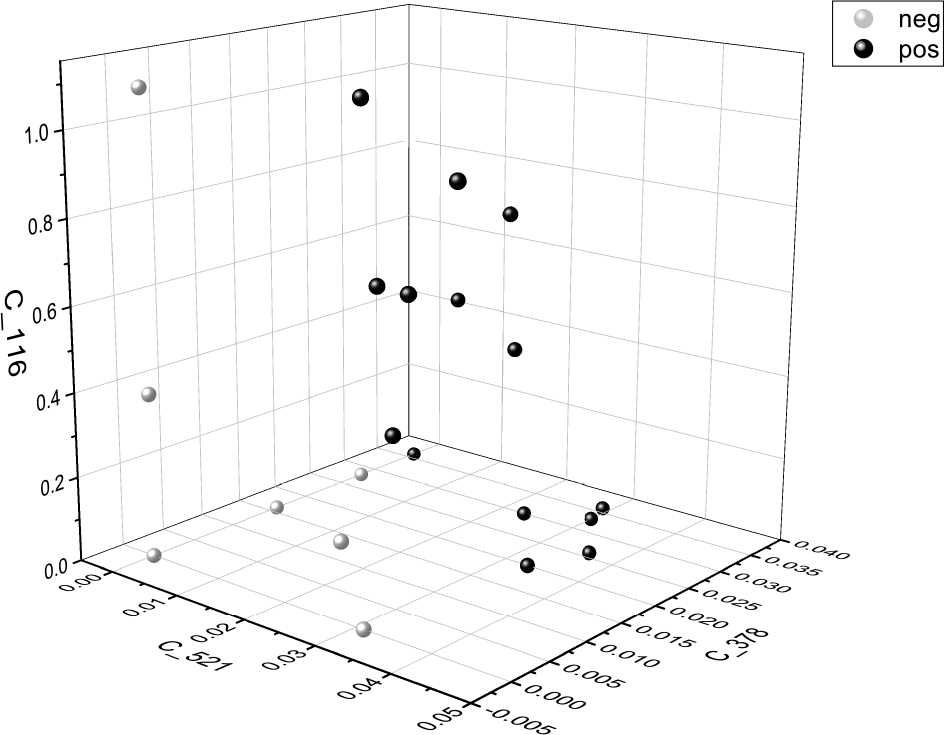
3D- scatterplot of signal intensities for cluster c_116, c_378 and c_521. White balls = Influenza-A negative, black balls = Influenza-A positive. The least significant cluster in the multivariate analysis is omitted, as it cannot be displayed in a 3D- diagram.

## 4. Discussion

To the best of our knowledge this is the first study indicating that Influenza-A-infections can be detected by aspirating nasal breath in infected patients.

There is only little known about breath analysis in viral diseases.

Most of the studies either have analyzed headspace over cell cultures [11,12] or the influence of vaccination on breath VOCs [13]. In another study dogs were used to detect the smell of virus infected cell cultures [14,15]. A recently published review compares different techniques for the detection of viral disease in breath [15].

There are studies using “breath biopsy” for the detection of cancer by differential ion mobility spectrometry but the device used in this study needs pre-analytic procedures with absorption/desorption tubes while using the STEP device the breath is aspirated during normal breathing and analyzed instantaneously [16]. This can also be performed by less trained staff and is well tolerated by the patients.

Another advantage is the speed of the analysis. Though using a retention time of 240 seconds most relevant VOCs could be found in the first 180 seconds. So, a reduction of the analysis time to 3 minutes seems to be possible.

Compared to PCR as the gold- standard, MCC-IMS is not only faster but does not need reagents and other supplies where shortages of swabs, especially flocked nasopharyngeal swabs and of less sensitive swabs have emerged in the ongoing SARS-CoV-2 pandemics.

Compared to gas chromatography coupled mass spectrometry the device is quite small, easily portable and can be used as a point-of-care diagnostic.

PCR-analysis of nasopharyngeal swabs used for the detection of viral disease may be false negative if the replication site is in the distal airways or the lung parenchyma [17]. Using a deep exhalation procedure all respiratory compartments are accessible by breath analysis.

One advantage of breath analysis by MCC-IMS is the speed and the ease of application. There is no need for preanalytics, sophisticated training of staff and no need of reagents or consumables that can be subject to shortages in epidemics and pandemics. As the device is drawing the samples by an internal pump the only task the staff has to perform is to place the catheter into one nostril. We assume that the analysis time can be reduced to less than 3 minutes which enables the method to be used for mass screening at airports or mass sports events.

One shortcoming of our study is the sample size. Due to the late start of the study and the weak Influenza season 2020 only few patients could be analyzed and no validation cohort was available.

It may be argued that the difference in VOCs between the groups is a result of inflammatory processes and not specific for Influenza-A.

As the 2020 Influenza epidemic faded out very quickly in the beginning of March due to the preventive measures in the upcoming COVID-19 pandemic, the study was extended to also investigate SARS-CoV-2 infected patients. In this extended study we could demonstrate that not only patients with Influenza-A can be discriminated from controls but from SARS-CoV-2 as well. So, a specific detection of VOCs in Influenza-A seems to be achievable.

After demonstrating the proof-of-principle for the MCC-IMS diagnostic in respiratory infections in the current study, future studies shall address the optimization of sampling procedure and analysis algorithms as well as extension to other viral infections. Besides statistical methods we are planning to implement methods of artificial intelligence.

Currently, we are working on the development of a MCC-IMS device for screening purposes.

In two recent studies posted as preprints MCC-IMS could also identify SARS-CoV-2 infections in breath. ^18,19^ As MCC-IMS is fast, non-invasive and ressource-saving it could be a breakthrough for SARS-CoV-2 mass screening for travellers and huge cultural events. Hence, we are urgently recommending to spur the further development of this new method.

## 5. Conclusion

To cut infection chains effective screening for Influenza-A and other communicable virus diseases is urgently needed.

In this proof-of-concept study MCC-IMS was able to discriminate Influenza-A-infections by simple aspiration of nasal breath during normal breathing. As the whole analysis takes less than 5 minutes MCC-IMS allows for rapid screening which is highly demanded in viral epidemics or pandemics especially in the scope of mass screening.

Further investigation of this method is needed as well as application to other viral diseases.

## Data Availability

data available on request

## Acknowledgements

The authors state that there is no conflict of interests.

There was no external funding Authors Contribution

CS Conceptualization, Data curation, Formal Analysis, Methodology, Investigation, Writing – original draft, Writing – review & editing

IS Visualization, Formal Analysis, Writing – original draft, Writing – review & editing

TB Investigation, Writing – review & editing

WS Writing – review & editing

The paper has been **accepted** as preprint to medrxiv.org **#**MEDRXIV/2020/099259

